# The divergent effects of nicotinamide riboside and high-intensity exercise training on skeletal muscle epigenetic aging

**DOI:** 10.64898/2026.01.16.26344093

**Authors:** Aino Heikkinen, Liina Uusitalo-Kylmälä, Ida Blom, Jørn Wulff Helge, Linn Gillberg, Robert Seaborne, Steen Larsen, Macsue Jacques, Robin Grolaux, Sari Aaltonen, Jaakko Kaprio, Birgitta W van der Kolk, Sini Heinonen, Nir Eynon, Kirsi H Pietiläinen, Riikka Kivelä, Eija Pirinen, Miina Ollikainen

**Author notes:** **Corresponding author:** Aino Heikkinen.

## Abstract

Aging is accompanied by a decline in physiological function and increased vulnerability to disease, with mitochondrial dysfunction and epigenetic alterations recognized as key hallmarks. Nicotinamide riboside (NR), a vitamin B3 precursor to NAD⁺, and high-intensity interval training (HIIT) have both been proposed to ameliorate aging-related mitochondrial decline, but their effects on skeletal muscle epigenetic aging are not fully elucidated. Here, we assessed the impact of 5-month NR supplementation and 4-6 weeks HIIT on epigenetic age acceleration (EAA, via seven epigenetic clocks) in human skeletal muscle across three independent studies. NR supplementation was associated with reduced muscle EAA, particularly when measured with the PCHannum, MEAT, and DunedinPACE clocks, while HIIT produced opposite effects in some clocks, notably increasing pace of aging by DunedinPACE. Correlation analyses revealed that changes in skeletal muscle mitochondrial content correlated with changes in MEAT-derived EAA after NR and 6-weeks of HIIT. Together, these findings indicate that skeletal muscle epigenetic aging can be modulated by NR and HIIT interventions but in opposing directions, highlighting a potential link between mitochondria abundance and epigenetic clocks. Further studies are warranted to clarify how NR and exercise regulate epigenetic aging. These results offer new insights into development of strategies for promoting epigenetic outcomes and healthy aging.

## 1 Introduction

Human aging is characterized by the progressive accumulation of cellular damage, leading to functional decline across tissues, increased disease burden, and ultimately death. Although aging affects all organs, skeletal muscle is particularly critical due to its role in mobility, metabolic health, and quality of life. Age-related deterioration of skeletal muscle contributes to sarcopenia and frailty, markedly increasing the risk of falls, disability, and mortality in older adults (Xu et al. 2021). Many hallmarks of aging are tightly intertwined with cellular metabolism (López-Otín et al. 2023; Kroemer et al. 2025), in which the mitochondrion is a central organelle known for converting chemical energy into ATP. Unsurprisingly, mitochondrial dysfunction is recognized as one of the hallmarks of aging. Aged skeletal muscle show lower mtDNA abundance, citrate synthase activity and diminished expression of mitochondrial related genes (Short et al. 2005; Murgia et al. 2017). These alterations, only partially explaind by decreased physical activity (Carter et al. 2015), are further linked to age-related changes such as chronic inflammation, oxidative stress, and impaired nutrient signalling. Collectively, declining mitochondrial content in skeletal muscle represents a key mechanism linking cellular aging to impaired muscle function.

A crucial regulator of cellular redox status is the coenzyme NAD^+^, which is essential for mitochondrial reactions including the oxidative phosphorylation and serves as a central cosubstrate for three major NAD^+^ consuming protein families: sirtuins, PARPs, and CD38. These enzymes are involved in processes like epigenetic regulation, mitochondrial metabolism, inflammation and oxidative stress —thereby linking NAD⁺ metabolism to multiple hallmarks of aging (Kroemer et al. 2025). Evidence suggests that NAD^+^ levels may decline with age in some tissues, including blood and skeletal muscle (Chini et al. 2017; Camacho-Pereira et al. 2016; Covarrubias et al. 2021), prompting interest in whether strategies to restore NAD^+^ levels could extend healthspan and delay onset of age-associated diseases. Two primary strategies have emerged to boost NAD^+^ levels: pharmacological supplementation with, for instance, NAD^+^ precursors and lifestyle interventions such as exercise training (Covarrubias et al. 2021).

Among NAD^+^ precursor vitamin B3s, nicotinamide riboside (NR) has been shown to be safe in humans and effective in increasing NAD^+^ metabolome both in blood (Airhart et al. 2017; Martens et al. 2018; Lapatto et al. 2023) and in skeletal muscle (Elhassan et al. 2019). In rodent models, NR supplementation improves skeletal muscle mitochondrial content and respiration, prevents high-fat diet-induced obesity, mitigates age-related muscle dysfunction and extends lifespan (Cantó et al. 2012; Lozada-Fernández et al. 2022; Li et al. 2023; Zhang et al. 2016; Seldeen et al. 2021; Frederick et al. 2016). However, the findings are mixed in humans, particularly regarding whole-body metabolic outcomes. Some studies have reported modest improvements in fat-free mass (Remie et al. 2020) and blood pressure (Martens et al. 2018) while others have found no effects on adiposity or insulin sensitivity (Dollerup et al. 2018; Martens et al. 2018; Lapatto et al. 2023). Furthermore, the effect of NR in skeletal muscle function remains understudied. Notably, we have previously shown that long-term (5-month) NR supplementation increases skeletal muscle mitochondrial content, decreases global DNA methylation and modifies methylation of CpG sites in mitochondria-related genes in monozygotic twins (Lapatto et al. 2023). Overall, while promising effects of NR have been observed, the evidence remains inconsistent—likely due to differences in intervention duration, participant characteristics, and specific outcome measures.

Exercise training is another promising booster of NAD^+^ metabolism (Walzik et al. 2023). Both aerobic and resistance exercise training have been shown to increase muscle NAD^+^ levels and the expression of muscle nicotinamide phosphoribosyltransferase (*NAMPT*) (de Guia et al. 2019; Costford et al. 2009; Lamb et al. 2020), the primary rate-limiting enzyme in the NAD^+^ biosynthesis in muscle. Exercise also induces broad metabolic benefits, including improved mitochondrial function and insulin sensitivity (Cartee et al. 2016; Irving et al. 2015). High-intensity interval training (HIIT) is a time-efficient exercise modality that improves cardiorespiratory fitness and metabolic health faster than traditional endurance training (Ko et al. 2025). HIIT can increase mitochondrial content in skeletal muscle (Mølmen et al. 2025) and alter muscle fiber type composition (Van der Stede et al. 2024). HIIT has also been shown to increase circulating NAMPT levels in blood (Walzik et al. 2025). Moreover, HIIT and other exercise forms can induce DNA methylation changes associated with a younger epigenetic profile (Ostaíza et al. 2025; Voisin et al. 2024).

Despite these advances, it remains unclear whether NR supplementation or HIIT can slow epigenetic aging as measured by epigenetic clocks—DNA methylation–based biological aging biomarkers associated with functional decline, disease, and mortality risk. While observational studies suggest that higher physical activity is associated with younger DNA methylation profiles (You et al. 2025; Ammous et al. 2025; Voisin et al. 2024), the evidence for HIIT specifically is limited. Furthermore, no studies to date have demonstrated similar effects for NR supplementation. Understanding whether these interventions can meaningfully influence epigenetic aging is essential, as epigenetic clocks offer potetially cost-effective tools for estimating biological age and evaluating anti-aging strategies. Addressing this gap is critical for optimizing interventions aimed at promoting healthy aging.

Here, we investigated whether 5-month NR supplementation in the Finnish Twin Cohort and HIIT interventions in the EpiH and Gene SMART studies influence epigenetic aging in skeletal muscle using seven epigenetic clocks. We additionally assessed the impact of NR on blood epigenetic aging as blood is the primary tissue used in epigenetic aging research and the main target for most epigenetic clocks. We also examined whether the potential intervention-induced changes in muscle epigenetic aging were associated with increases in mitochondrial content. Our findings suggest that NR and HIIT may both modulate epigenetic aging, often in opposing directions, with responses varying by individual and epigenetic clocks. These results encourage further research to determine how NR supplementation and exercise modalities can be leveraged to promote healthy aging, while accounting for individual variability and differences in epigenetic clocks.

## 2 Materials & Methods

### 2.1 Study Cohorts

#### 2.1.1 Long-term human NR trial

The human NR trial was originally performed to investigate the effect of long-term 5-month NR supplementation (with the dose of 1000mg/day for 5 months) on muscle and adipose tissue mitochondrial biogenesis (clinicaltrials.gov entry NCT03951285) using monozygotic twin pairs, targeting twin pairs discordant for body mass index (BMI). These pairs originate from three larger longitudinal Finnish Twin Cohorts (FTC; FinnTwin12 (Rose et al. 2019), FinnTwin16 (Kaidesoja et al. 2019) and Older Finnish Twin Cohort (Kaprio et al. 2019)). The full study design, and exclusion and inclusion criteria are described in detail in the original study paper (Lapatto et al. 2023).

In the current study, we utilized data on adult twin individuals (30–65 years) who were administered NR supplementation (1000 mg/day for 5 months) with available DNA methylation samples from both timepoints (before and after NR) from either of the two tissues: blood (n=36 individuals) and muscle (n=30 individuals). We included twin individuals without any data from their co-twin to increase the number of samples in each analysis that was performed at the individual-level. For the comparison between the influence of NR on epigenetic ages between leaner and heavier co-twins, we included only complete BMI-discordant twin pairs (n=14 and 15 pairs in muscle and blood, respectively) defined as having a minimum difference of 2.5 BMI units between co-twins.

Venous blood and skeletal muscle samples (*vastus lateralis*) were collected from each participant at baseline and after 5-months of NR supplementation (Lapatto et al. 2023). The muscle biopsies were collected using a Bergström needle under local anesthesia, snap-frozen and stored in liquid nitrogen, and then transferred to -80 °C. The blood samples were stored at -80 °C. High-molecular weight DNA was extracted from muscle and blood samples using QIAmp DNA Mini kit (Qiagen) and bisulfite-converted by an EZ DNA Methylation kit (ZYMO Research) for the quantification of DNA methylation levels.

#### 2.1.2 EpiH study

The EpiH samples are derived from an exercise training intervention study in which 20 younger (21-42 years) and 20 older (55-74 years) individuals with overweight or obesity (BMI range 27-44 kg/m²) were recruited (Søgaard et al. 2018). The inclusion criteria as well as the detailed study protocol are described elsewhere (Søgaard et al. 2018; Søgaard et al. 2019). Briefly, the individuals completed a six-week supervised HIIT intervention three times a week on a bicycle ergometer. Each HIIT session consisted of two minutes warm up followed by five one-minute-intervals at 124 ± 12% of their maximal load (W) interrupted by 90 seconds of cycling at 25 W or just resting on the bike. After two weeks of training, the load was increased by 10%. An incremental test on a bicycle ergometer was performed before and after the intervention to determine maximal oxygen uptake during exercise (VO_2max_) which reflects cardiorespiratory fitness. Two VO_2max_ tests were performed on two separate test days before the HIIT intervention to avoid potential learning effects.

Muscle biopsies from the *vastus lateralis* were collected prior to (maximum one week) and after the intervention (48-72 h after the final HIIT session) using a Bergström needle and manual suction in an overnight fasted condition (Søgaard et al. 2018). Muscle tissue was immediately snap-frozen in liquid nitrogen and thereafter stored at -80°C. The frozen tissue was treated with 2 mg/mL (0.2%) collagenase (Clostridium histolyticum, Type I, Sigma-Aldrich) and myofibers were extracted manually to enrich myofibers and to remove non-myofiber material. Genomic DNA was extracted from the washed and frozen down pellet of sorted myofibers using the DNeasy Blood and Tissue Kit (Qiagen, Maryland, USA).

#### 2.1.3 Gene SMART study

The Gene SMART (Skeletal Muscle Adaptive Response to Training) study is a multi-centre exercise training study aiming to identify OMIC biomarkers of the response to 4 weeks of HIIT (Jacques et al. 2020). To date, 100 male and 30 female young (18-48 years old), healthy, moderately-fit participants have undersgone four-week supervised HIIT intervention on an electronically braked cycle ergometer. The detailed Gene SMART methods and study design can be found elsewhere (Yan et al. 2017; Jacques et al. 2023). Briefly, the 4-week HIIT program included three supervised sessions per week. Each session was preceded by a 5-minute warm-up at 50W, followed by six to twelve two-minute intervals at intensities ranging progressively from 40% to 70% above the individual’s lactate threshold, interspersed with 1-minute recovery periods. VO₂_max_ was assessed before and after four intervention using a protocol consisting of 2-minute exercise bouts performed to exhaustion.

Muscle samples were collected from the *vastus lateralis* before and after (48 hours after the last session) HIIT intervention using a Bergström needle and manual suction.The samples were frozen in liquid nitrogen and stored at −80°. Approximately 15 mg of homogenized bulk muscle tissue was used for DNA extraction with the Qiagen AllPrep DNA/RNA kit.

### 2.2 Myotube cell culture

Primary human myoblasts were plated into 6-well plates (∼120,000 cells / well) and when the cells reached 80-90% confluency (typically within 2-4 days), a differentiation medium was introduced to promote maturation into myotubes. Cells were cultured in the differentiation medium for 6 days. Following the differentiation, the cells were exposed to a medium containing 1mM of NR (Novalix Pharma, Strasbourg, France) (n=6 technical replicates) or a control medium without NR (n=6 technical replicates). A concentration of 1 mM NR has been shown to produce the maximal increase in intracellular NAD⁺ levels in C2C12 myotubes (Cantó et al. 2012) and is typically the highest concentration used in *in vitro* experiments. Due to the rapid degradation of NR, the medium was replaced daily. After 72 hours of exposure, cells were harvested for analysis. The contents of the cultivating media are described in Supplementary Materials.

For DNA extraction, we used the AllPrep DNA/RNA/miRNA Universal Kit (Qiagen) according to the manufacturer’s instructions, with the modification that DNA was eluted using 2 x 50 µL of RNase-free water. To measure NAD^+^ levels, cells were first washed with PBS. Subsequently, three wells were pooled together into an Eppendorf tube, and the cell pellet was collected following centrifugation at 1400 rpm for 10 minutes. The quantification of NAD^+^ levels was performed using the NADMED assay (Euro et al. 2025).

### 2.3 Epigenetic ages

#### 2.3.1 DNA methylation data preprocessing

DNA methylation data (.idat files) from skeletal muscle across all studies (human NR trial, myotube cell culture, the EpiH cohort, and the Gene SMART cohort; GEO accession: **GSE171140**) and from blood in the NR trial only, were normalized and preprocessed using the *meffil* R package (Min et al. 2018) following a pipeline described in detail elsewere (Heikkinen et al. 2025). Methylation data from the human NR trial were generated using the Illumina Infinium MethylationEPIC v1 array, while all other DNA methylation data were generated using the EPIC v2 array.

#### 2.3.2 Epigenetic clocks

Epigenetic age estimates were calculated from the preprocessed DNA methylation beta values using PC-based versions (Higgins-Chen et al. 2022) of the original Hannum (Hannum et al. 2013), Horvath (Horvath 2013), PhenoAge (Levine et al. 2018), and GrimAge clocks (Lu et al. 2019). In addition, we computed the pace of aging measure DunedinPACE (Belsky et al. 2022), GrimAge2 (Lu et al. 2022), as well as muscle-specific MEAT clock for DNA methylation data derived from muscle tissue (Voisin et al. 2020). Both PCGrimAge and the more recent GrimAge2 were included since the PC-based version may reduce susceptibility to technical noise whereas GrimAge2, still lacking a PC-based version, incorporates two additional plasma-based biomarkers in the prediction. In addition, updated MEAT clock, the MEAT2 (Voisin et al. 2021), was used for all other studies except for the Gene SMART, which was used in the development of MEAT2, and therefore the original MEAT1 (Voisin et al. 2020) was applied instead. Epigenetic age acceleration (EAA) was defined as the residuals from regressing epigenetic age estimates on chronological age and was calculated separately for each study. Outliers, defined as values exceeding ±3 standard deviations from the study mean EAA, were excluded from downstream analyses. In the NR trial, one outlier was identified for muscle (GrimAge2) and three for blood (one in DunedinPACE, two in GrimAge2). In EpiH, there were no outliers whereas in Gene SMART, two outliers were detected with the GrimAge2 clock.

### 2.4 Mitochondrial content

Relative mitochondrial DNA quantity (mtDNAq) was assessed in the NR trial muscle biopsies using quantitative PCR (qPCR) from phenol-chloroform extracted DNA. The mtDNA amount was determined by quantifying three mitochondrial genes—*D-Loop*, *cytochrome b* (*CYTB*) and *16S rRNA* relative to three nuclear reference genes: *amyloid beta Precursor Protein* (APP), *Beta-2-Microglobulin* (B2M) and *Hemoglobin Subunit Beta* (HBB). The data was processed using qbase+ software (Biogazelle) by standard curve method. The average calibrated normalized relative quantities (CNRQs) of the mitochondrial target genes were used to calculate mtDNAq for each sample.

In cultured myotubes, mtDNAq was similarly measured using qPCR, targeting the mitochondrial genes *NADH Dehydrogenase Subunit 5* (ND5) and CYTB, normalized to the nuclear genes APP and B2M. The mean of the ND5 and CYTB CNRQ measures was used to represent mtDNAq for each sample.

In HIIT studies, measurements of the citrate synthase (CS) activity were used as markers for the mitochondrial content in the skeletal muscle. In EpiH, the CS activity was calculated using spectrophotometry from 2mg freeze-dried and dissected muscle tissue as described elsewhere (Frandsen et al. 2022). In Gene SMART, small pieces of freeze-dried skeletal muscle tissue were lysed in an ice-cold buffer (KH_2_PO_4_ & K_2_HPO_4_) and total CS activity was measured in triplicates using spectrophotometric assays (Jacques et al. 2021).

### 2.5 Statistical analysis

All the analyses were perfomed in R statistical software (version 4.5.1).

#### 2.5.1 Perfomance of epigenetic clocks in predicting chronological age

We used Pearson correlation to evaluate how well epigenetic clocks that provide age estimates in years—excluding DunedinPACE—predict chronological age. To quantify the average deviation between predicted and actual age, we calculated the mean absolute error (MAE).

#### 2.5.2 Before versus after intervention

Differences in EAA before and after NR supplementation or HIIT were assessed using linear mixed-effects models (*lmerTest* R package) adjusted for chronological age, sex, and BMI. In the human NR trial, models were additionally adjusted for self-reported smoking status. To account for repeated measures, models included random effects: unique participant identifiers for the HIIT studies, and participant ID nested within family ID in the NR trial to account for both repeated measures and relatedness of twins in a pair.

In the myotube experiment, differences in EAA between control and NR-treated cells were analyzed using linear models. Changes in NAD⁺ levels and mtDNAq were assessed using the Wilcoxon signed-rank test.

To investigate whether leaner and heavier co-twins in BMI-discordant twin pairs differed in EAA changes following NR, we calculated pre vs post EAA changes and used linear mixed effects models to assess the effect of weight status (i.e., lean or heavy), adjusting for sex, smoking and family ID.

#### 2.5.3 Intra-class correlation

The intra-class correlation coefficients (ICCs) were estimated using linear mixed-effects models with a random intercept for family ID, with either the baseline EAA or change in EAA upon as an outcome. For the analysis of change in EAA, mean baseline EAA within a twin-pair was included as a covariate to estimate within-family resemblance in change of EAA upon NR independent of baseline values. The 95% confidence intervals for the ICC estimates were obtained using parametric bootstrapping with the *bootMer* function in R, based on 1,000 simulated datasets drawn from the fitted models.

#### 2.5.4 Correlation analyses

The correlations between delta variabes (i.e., post-intervention minus pre-intervention values within an individual) of different EAA clocks and mtDNAq in the NR trial, and of different EAA clocks, CS activity and and VO₂_max_ in EpiH and Gene SMART studies were estimated using Pearson correlation. Additionally, in the NR trial, the correlation between EAA and mtDNAq within twin pairs was assessed using Pearson correlation on the difference in changes between co-twins (i.e., delta-delta variables, calculated by subtracting the change in twin 1 from the change in twin 2).

## 3 Results

Figure 1 illustrates the study design, and Table 1 summarizes the participant characteristics of the three human studies. Mean ages ranged from 33 years in Gene SMART to 40 in NR study and 48 in EpiH, while female participants comprised 44-48%. The mean baseline BMI was 24.4 kg/m²in the Gene SMART, whereas NR and EpiH studies included predominantly individuals with overweight or obesity (mean baseline BMIs 30.3 and 32.0 kg/m², respectively).

**Figure 1.**
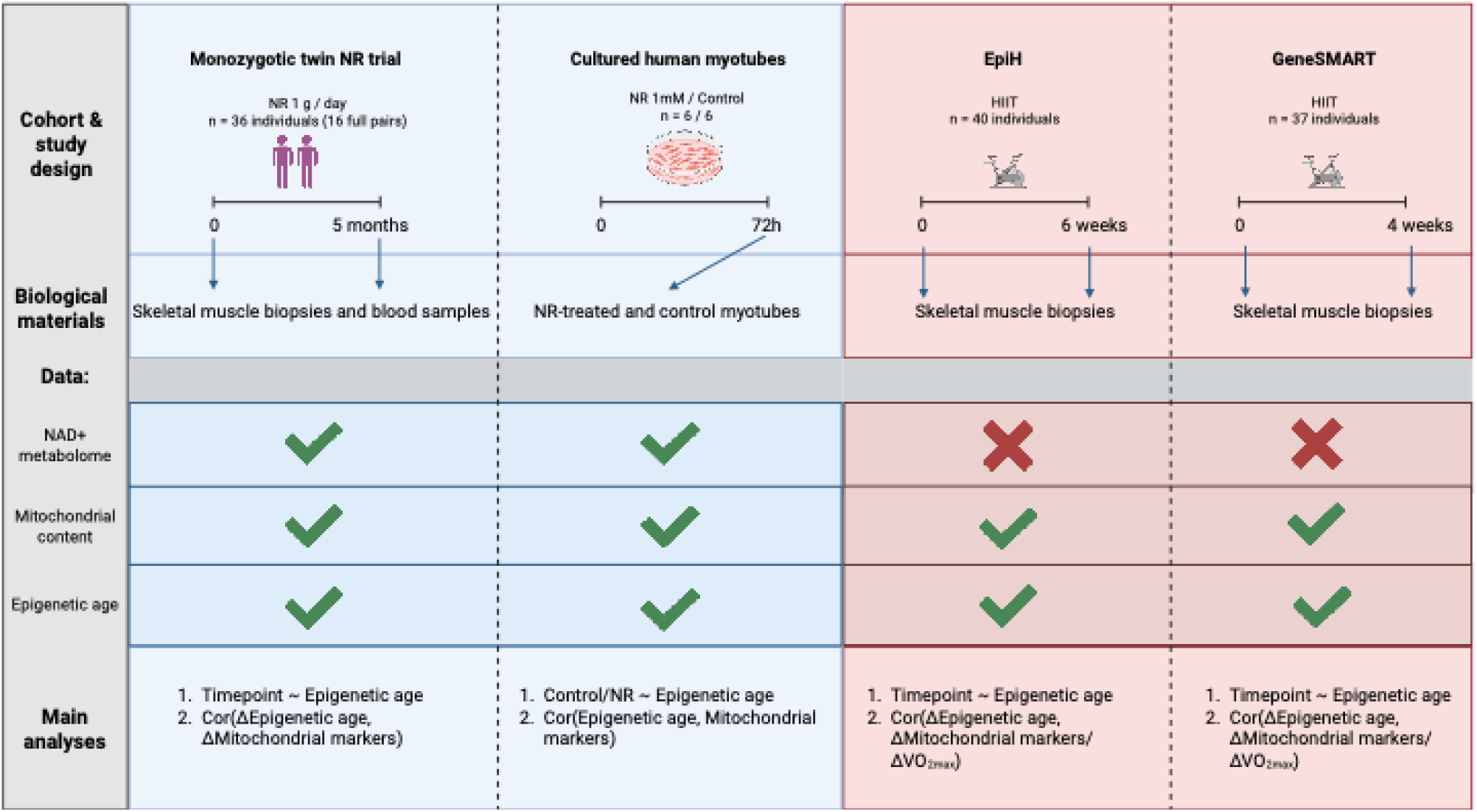
Schematic representation of the study design, datasets, and statistical analysis for each study. Created with Biorender.com.

**Table 1.**
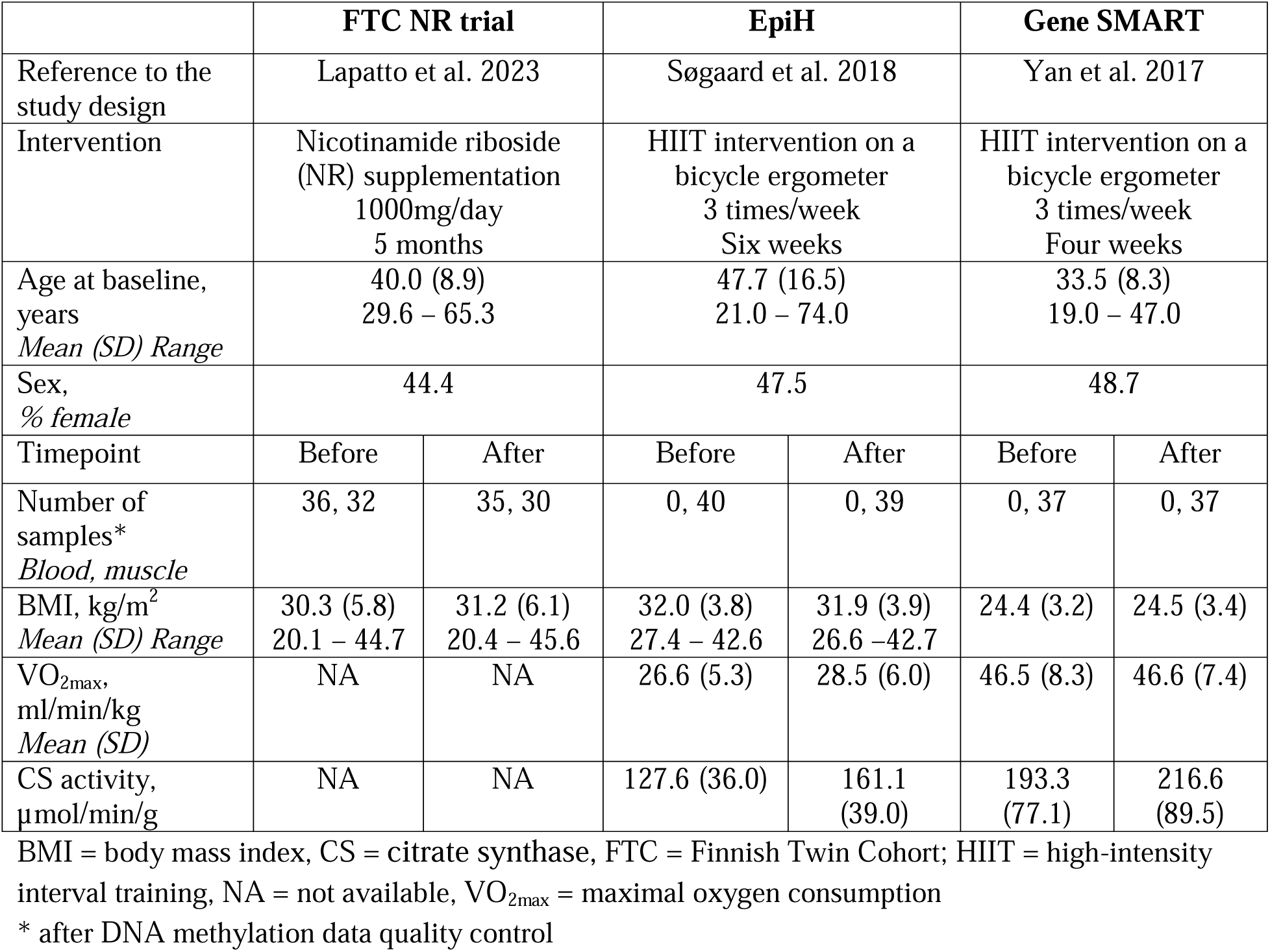
Characteristics of participants included in this study.

|  | FTC NR trial |  | EpiH |  | Gene SMART |  |
| --- | --- | --- | --- | --- | --- | --- |
| Reference to the study design | Lapatto et al. 2023 |  | Søgaard et al. 2018 |  | Yan et al. 2017 |  |
| Intervention | Nicotinamide riboside (NR) supplementation<br>1000mg/day<br>5 months |  | HIIT intervention on a bicycle ergometer<br>3 times/week<br>Six weeks |  | HIIT intervention on a bicycle ergometer<br>3 times/week<br>Four weeks |  |
| Age at baseline, years<br>*Mean (SD) Range* | 40.0 (8.9)<br>29.6 – 65.3 |  | 47.7 (16.5)<br>21.0 – 74.0 |  | 33.5 (8.3)<br>19.0 – 47.0 |  |
| Sex, % female | 44.4 |  | 47.5 |  | 48.7 |  |
| Timepoint | Before | After | Before | After | Before | After |
| Number of samples*<br>*Blood, muscle* | 36, 32 | 35, 30 | 0, 40 | 0, 39 | 0, 37 | 0, 37 |
| BMI, kg/m<sup>2</sup><br>*Mean (SD) Range* | 30.3 (5.8)<br>20.1 – 44.7 | 31.2 (6.1)<br>20.4 – 45.6 | 32.0 (3.8)<br>27.4 – 42.6 | 31.9 (3.9)<br>26.6 – 42.7 | 24.4 (3.2) | 24.5 (3.4) |
| VO<sub>2max</sub>, ml/min/kg<br>*Mean (SD)* | NA | NA | 26.6 (5.3) | 28.5 (6.0) | 46.5 (8.3) | 46.6 (7.4) |
| CS activity, μmol/min/g | NA | NA | 127.6 (36.0) | 161.1 (39.0) | 193.3 (77.1) | 216.6 (89.5) |

BMI = body mass index, CS = citrate synthase, FTC = Finnish Twin Cohort; HIIT = high-intensity interval training, NA = not available, VO<sub>2max</sub> = maximal oxygen consumption
\* after DNA methylation data quality control

### 3.1 Performance of the epigenetic clocks

Because the epigenetic clocks—except the muscle-specific MEAT clock—were originally developed for blood tissue but are applied here to both blood and skeletal muscle, we first assessed their ability to capture age-associated DNA methylation changes in blood (NR study) and in skeletal muscle (NR, EpiH, and Gene SMART studies). DunedinPACE, which estimates the pace of aging (years per calendar year) rather than epigenetic age, was excluded from this analysis.

In blood, all epigenetic clocks showed strong correlations with chronological age, as expected, with correlation coefficients ranging from R² = 0.80 to 0.93 (Supplementary Table 1). However, some clocks exhibited relatively high median absolute errors (MAE), which is an estimate of error between paired measurements, most notably PCGrimAge, with MAE values reaching up to 13.6 years.

In muscle, the muscle-tissue specific MEAT clock performed well across all three studies (NR trial, EpiH, and Gene SMART), showing both strong correlations and low MAE values with chronological age (Supplementary Table 1). GrimAge2 also effectively captured age-related methylation changes in muscle, with high correlation (R² = 0.90 – 0.98) and moderate MAE (5.9 – 10.5). While PCGrimAge also correlated strongly with chronological age in muscle, its MAE ranged from 24.8 to 30.6 years, highlighting substantial calibration issues, i.e., that the clock underperforms in estimating chronological age in these samples. The weakest performance was observed for PCHorvath, PCHannum, and PCPhenoAge, with maximum correlation coefficients around R² = 0.70 in the NR trial and EpiH studies, and R² = 0.50 in Gene SMART. Corresponding MAE values varied widely, from 9.8 years (NR PCHorvath) to 35.5 years (Gene SMART PCPhenoAge), again indicating poor calibration.

Overall, differences in MAEs across clocks reflect variability in their ability to estimate age in years, specifically in skeletal muscle, while most still capture some age-related epigenetic changes as indicated by their moderate-to-high correlations with chronological age.

### 3.2 Impact of 5-month NR supplementation on epigenetic age acceleration in human blood and skeletal muscle

To investigate the potential rejuvenating effects of long-term NR supplementation, we determined whether the 5-month NR supplementation was associated with differences in the EAA measures in both blood and skeletal muscle tissues. In blood tissue (n=36 individuals), we observed a significant decrease in DunedinPACE, PCHorvath and PCHannum after 5-months of NR supplementation (Supplementary Table 2). In muscle tissue (n=30 individuals), we saw that, on average, the mean EAA was lower after NR in all clocks but PCGrimAge showed an increase in EAA (ß=0.52, p=0.007) (Figure 2A, Table 2). However, the decrease was statistically significant only when measured with DunedinPACE (ß=-0.04, p<0.001), PCHannum (ß=-0.69, p=0.013) and MEAT (ß=-2.39, p<0.001) (Figure 2A). To investigate whether the observed changes in muscle epigenetic aging could be replicated *in vitro* setting with a placebo control, primary human myotubes were treated with NR or vehicle. NR exposure significantly increased intracellular NAD⁺ levels (1.52-fold compared with control cells; Supplementary Figure 1A). Mitochondrial DNA content was slightly higher in NR-exposed cells but did not reach statistical significance (Supplementary Figure 1B). No EAA measures showed a statistically significant difference between NR-treated and control cells (Supplementary Figure 1C), although the MEAT clock indicated a marginally higher epigenetic age in NR-exposed cells (β = 1.55, p = 0.05). Notably, all four clocks showed effect directions opposite to those observed in the human trial.

**Figure 2.**
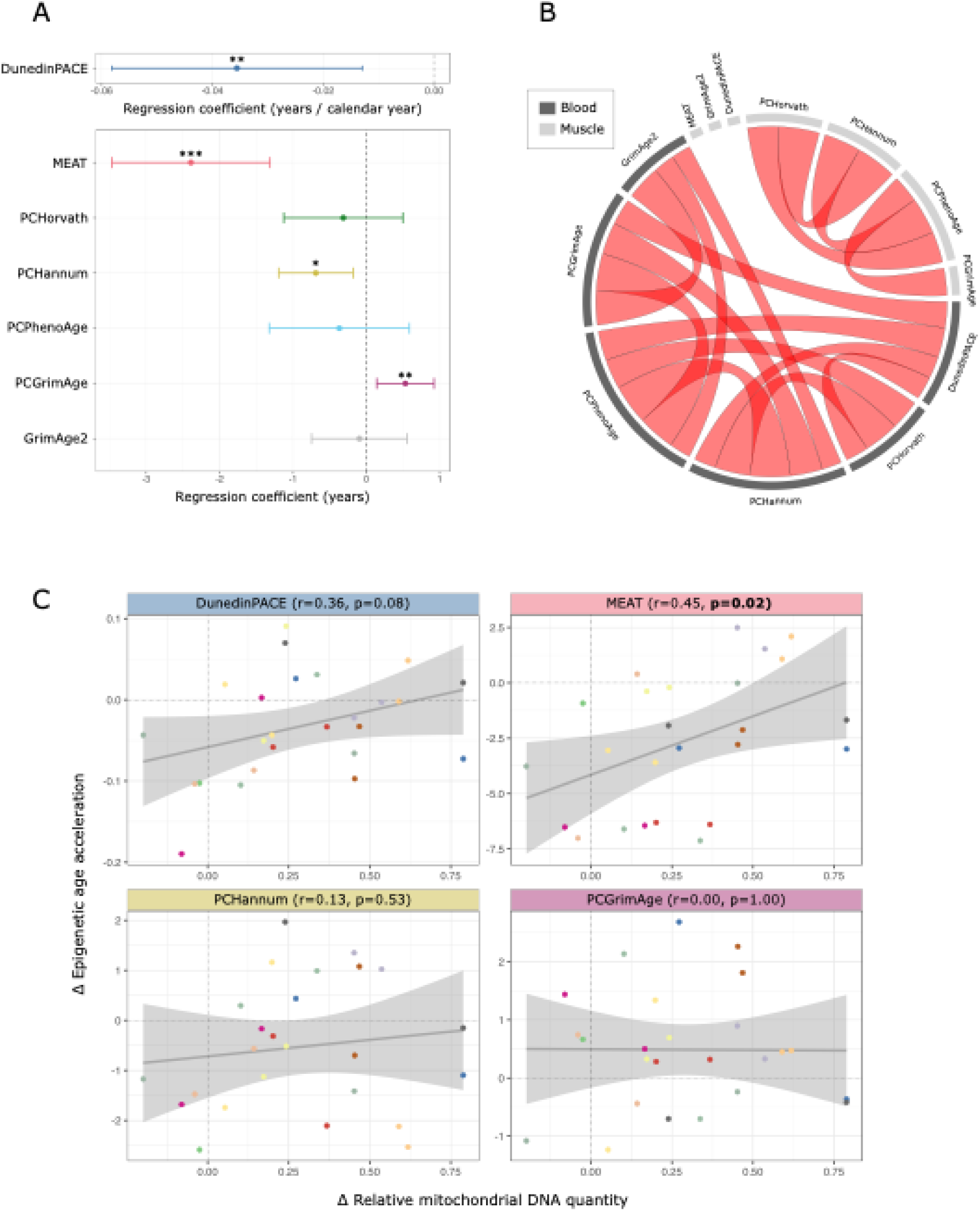
The impact of nicotinamide riboside (NR) treatment in human skeletal muscle epigenetic age acceleration (EAA). A) Forest plots showing the effect of 5-month NR supplementation in human skeletal muscle (n=30 individuals). P-values are derived from linear mixed effect models. B) Circosplot depicting the correlations between changes in different EAA estimates after 5-month NR supplementation derived either from blood (dark grey) or muscle tissues (light grey) within the same individual (n=29 individuals). Only p<0.05 correlations are shown. All observed correlations were positive. C) Correlation between changes in EAA measures and corresponding changes in mitochondrial DNA quantity in skeletal muscle following 5-month NR supplementation (n=30 individuals). Each color represents a distinct twin pair. Δ = difference in post vs. pre values; r = Pearson correlation coefficient *p<0.05, **p<0.01, ***p<0.001

**Table 2.** Epigenetic age acceleration measures at baseline and after 5-month nicotinamide riboside (NR) supplementation (n=30 individuals), along with intra-class correlation coefficients (ICCs) (n=14 complete twin pairs) in muscle. P-values are derived from linear mixed-effects models adjusted for chronological age, sex, smoking, BMI, with personID nested within familyID as a random effect. P-values<0.05 are bolded.

| Clock | Mean (SD)* at baseline | Mean (SD)* at 5-month | Change (SD)* | P-value | ICC (95% CI) baseline | ICC (95% CI) change |
| --- | --- | --- | --- | --- | --- | --- |
| DunedinPACE | 1.38 (0.07) | 1.34 (0.06) | -0.04 (0.06) | **0.001** | 0.78 (0.4-0.92) | - |
| PCHorvath | 0.16 (2.44) | 0.03 (2.36) | -0.13 (2.00) | 0.321 | 0.41 (0-0.75) | 0.45 (0-0.78) |
| PCHannum | 0.27 (1.60) | -0.23 (1.48) | -0.50 (1.24) | **0.013** | 0.24 (0-0.66) | 0.01 (0-0.54) |
| PCPhenoAge | 0.15 (2.33) | -0.15 (2.45) | -0.30 (2.34) | 0.335 | 0.56 (0.07-0.84) | - |
| PCGrimAge | -0.25 (1.67) | 0.25 (1.39) | 0.50 (0.95) | **0.007** | 0.68 (0.23-0.89) | - |
| GrimAge2 | 0.12 (1.46) | 0.05 (1.59) | -0.12 (1.79) | 0.757 | 0.36 (0-0.73) | 0.08 (0-0.58) |
| MEAT | 0.84 (2.54) | -1.65 (3.04) | -2.49 (2.88) | **2.2E-04** | 0.66 (0.22-0.88) | 0.68 (0.21-0.88) |

\* Units are in years, except for DunedinPACE, which is expressed in years/calendar year.

Given that our NR cohort consisted mostly of BMI-discordant monozygotic twin pairs (n=15 and 14 complete twin pairs in blood and muscle, respectively), we further investigated whether the leaner versus heavier co-twin status influenced changes in EAA following NR in those estimates showing significant changes after NR. No significant differences in response to NR were observed between leaner and heavier co-twins in either blood or muscle tissue (Supplementary Table 3).

To quantify the relative contributions of between-pair and within-pair variation in EAA both at baseline and in response to NR supplementation, ICC coefficients were estimated. ICC reflect the degree of resemblance between co-twins, with higher values indicating greater similarity. At baseline, ICCs for blood-based EAA measures ranged from 0.86 for PCHorvath to 0.33 for PCGrimAge (Supplementary Table 2). In muscle tissue, baseline ICCs ranged from 0.78 for DunedinPACE to 0.24 for PCHannum (Table 2). The ICCs for changes in EAA following NR supplementation were generally lower than those observed at baseline. Interestingly, MEAT and PCHorvath in muscle showed comparable ICCs for change and baseline values.

To assess whether changes in different EAA estimates were correlated in response to NR, we leveraged multiple epigenetic clock measures from both blood and muscle tissues in the same individuals in the NR trial. We examined correlations between changes in EAA (delta variables, i.e., post-value minus pre-value) across clocks and tissues. Significant correlations were observed between clocks within the same tissue, but no cross-tissue correlations were detected (Figure 2B). Notably, the skeletal muscle MEAT, DunedinPACE, and GrimAge2 clocks were the only ones whose changes after NR were not correlated with any other EAA estimates.

Together, our results suggest that 5-month NR supplementation is generally associated with reduced blood and muscle epigenetic aging similarly between leaner and heavier co-twins, although the effects vary by both tissues and clocks.

### 3.3 Correlation between changes in epigenetic age acceleration and mitochondrial content in skeletal muscle after NR supplementation

Although, on average, both skeletal muscle mtDNAq (1.36 fold higher, p_lmerTest_<0.001) and some of the EAA estimates were altered following NR supplementation, there was considerable inter-individual variability in these changes. To explore whether changes in skeletal muscle mitochondrial content might be associated with shifts in epigenetic aging, we examined correlations between mtDNAq and EAA following NR supplementation, focusing on those epigenetic clocks that showed significant alterations after NR. The analysis was restricted to skeletal muscle, as we did not have mtDNAq measures from blood. Interestingly, we found a positive correlation between changes in MEAT EAA and mtDNAq (r=0.45, p=0.02; Figure 2C) after NR, indicating that individuals with a greater increase in muscle mtDNAq tended to exhibit a smaller reduction in MEAT epigenetic aging. Other clocks did not show statistically significant correlations, although DunedinPACE displayed a similar trend with marginal significance (r=0.36, p=0.08) (Figure 2C). In the *in vitro* myotube experiment, we observed no correlations between EAA and mtDNAq.

We also examined within-pair differences in changes in MEAT-derived EAA and mtDNAq to assess whether the associations persisted after accounting for shared genetic and environmental factors. Notably, MEAT showed a significant positive correlation between the within-pair differences in EAA change and mtDNAq change (r = 0.50, p = 0.013), indicating that this clock may be sensitive to changes in skeletal muscle mitochondrial content independent of the underlying genetic factors.

### 3.4 Impact of high-intensity exercise training on epigenetic age acceleration in skeletal muscle

Given that exercise training has been shown to increase the skeletal muscle NAD^+^ metabolome in humans (de Guia et al. 2019; Lamb et al. 2020), we explored whether HIIT interventions induced changes in muscle EAA using data from two studies, EpiH (n=39 individuals) and Gene SMART (n=37 individuals). In EpiH (6-week HIIT intervention), out of the seven clocks that were investigated, PCGrimAge showed a significant decrease in EAA after HIIT (ß=-0.26, p=0.005), and PCHannum and DunedinPACE showed an increase in EAA (ß=0.47, p=0.023 and ß=0.02, p=0.010, respectively) (Figure 3A). After a 4-week HIIT intervention (Gene SMART), the only significant change in EAA we observed was an increase in DunedinPACE (ß=0.02, p=0.034) before versus after the exercise intervention (Figure 3A) similarly to what observed in EpiH. Notably, the clocks that demonstrated a significant decrease in EAA upon NR, showed increased EAA after HIIT (DunedinPACE). In contrast, PCGrimAge demonstrated significantly higher EAA after NR and lower EAA after HIIT (Figures 2A, 3A). The muscle-specific MEAT clock did not change after HIIT. These results indicate that 4–6 weeks of HIIT may alter skeletal muscle epigenetic aging in directions opposite to those observed with NR supplementation in humans.

**Figure 3.**
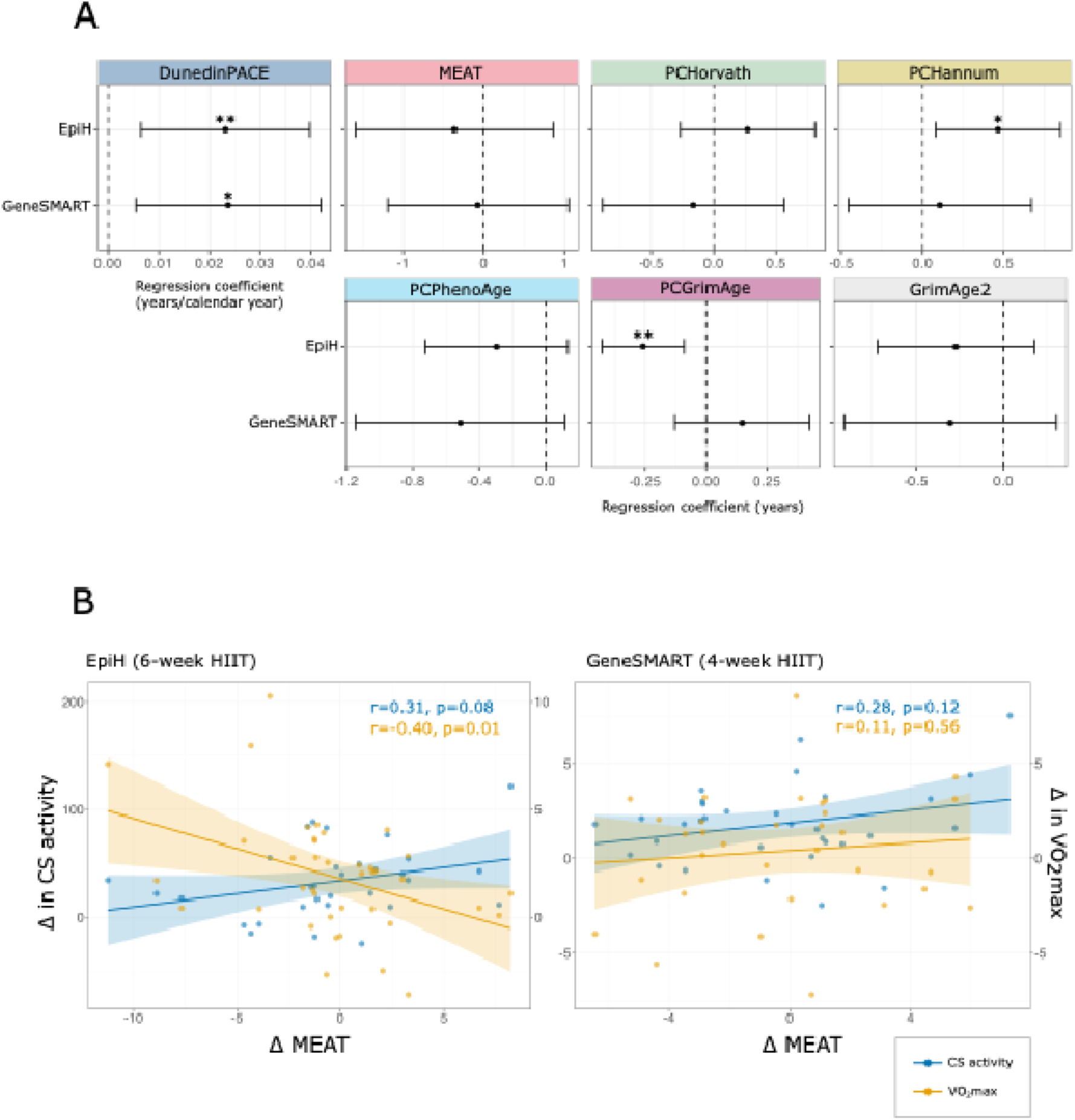
A) Forest plot illustrating the associations between high-intensity interval training (HIIT) and epigenetic age acceleration (EAA) across two studies. Effect sizes and 95% confidence intervals are shown. B) Correlation between changes in MEAT-derived EAA and changes in VO₂_max_ (in yellow) and citrate synthase (CS) activity (in blue) following HIIT in the EpiH and Gene SMART studies. EpiH n = 39 individuals, Gene SMART n = 37 individuals. Δ = difference in post vs. pre values; *p<0.05, **p<0.01

### 3.5 Correlation between changes in skeletal muscle epigenetic age acceleration, citrate synthase activity and cardiorespiratory fitness after high-intensity exercise training

Both 4-week Gene SMART and 6-weeks EpiH HIIT interventions has been previously shown to improve participants’ VO_2max_ and increase muscle specific mitochondrial citrate synthase (CS) activity, an estimate of mitochondrial content in human skeletal muscle (Larsen et al. 2012; Yan et al. 2017). We investigated whether the changes in EAA were correlated with corresponding changes in VO₂_max_ or CS activity in both HIIT studies. In the EpiH study, changes in MEAT following HIIT showed a negative correlation with changes in VO₂_max_, but a positive correlation with CS activity (Figure 3B). A similar positive trend between CS activity and MEAT was observed in the Gene SMART, although it did not reach statistical significance (Figure 3B). This positive association aligns with findings from the human NR trial, where MEAT changes were also positively associated with mtDNAq changes. In contrast, other epigenetic clocks demonstrated inconsistent associations across the two HIIT studies (Supplementary Figure 2).

To account for the broad age range in the EpiH study, we then stratified the analyses by age into younger (age 21-42 years, n=19) and older (age 55-74 years, n=20) subgroups. This allowed us to determine if the younger EpiH subgroup exhibited trends more comparable to the Gene SMART cohort (age 19-47). While stratification reduced the sample size and attenuated correlation coefficients, the direction of associations remained largely consistent with the primary analysis of entire EpiH study (Supplementary Table 4). Collectively, these results suggest that HIIT-induced changes in skeletal muscle mitochondrial content may be reflected in the MEAT clock, mirroring the pattern observed following NR supplementation.

## 4 Discussion

Given the aging population, it is essential to identify factors that promote healthy aging and biomarkers that best reflect and predict aging across tissues. Here, we use EAA as a biomarker of biological aging and show that both NR supplementation and HIIT – two well known mitochondrial content and NAD^+^ boosting strategies – may modulate skeletal muscle epigenetic aging. Notably, these interventions appeared to work in opposite directions: NR was generally associated with reduced EAA, while HIIT was associated with higher EAA. Furthemore, the changes in EAA, measured specifically by the muscle-specific MEAT clock showed significant positive correlations with changes in mitochondrial content across both intervention types. Collectively, these findings demonstrate the potential of epigenetic clocks as tissue-level biomarkers to evaluate modulators of biological aging, yet they also underscore the need to account for intervention- and clock-specific nuances when interpreting these effects on human aging.

While the effects of NR on human epigenetic aging has not been widely demonstrated before, one study has shown a reduction in epigenetic aging in blood following 12 week of NR supplementation in individuals with chronic obstructive pulmonary disease (Norheim et al. 2024). Furthermore, interventions known to modulate the NAD^+^ pathway and relate to lifespan improvements, including calorie restrication (CR) and enhanced diet quality, have also noticed decreased epigenetic aging post-interventions (Waziry et al. 2023; Fiorito et al. 2021; Fitzgerald et al. 2021; Maegawa et al. 2017). For instance, Waziry and collegues demonstrated that two years of CR led to a significant reduction in DunedinPACE (Waziry et al. 2023). Interestingly, we also observed a decrease in DunedinPACE in blood after 5-month of NR supplementation. Similarly to blood, DunedinPACE, along with MEAT and PCHannum, were significantly lower after NR in skeletal muscle. This finding contrasts a mice study that showed that life-long NAD^+^ depletion did not promote accelerated aging or mitochondrial defects (Chubanava et al. 2025). However, they did not assess the impacts on EAA in restoring or enhancing the NAD^+^ levels. Although we did not observe any measurable improvements in conventional metabolic parameters after 5-month NR (Lapatto et al. 2023), the observed reduction in some of the EAA markers may indicate some early molecular adaptations that precede systemic effects. The mechanisms underlying these changes remain unclear, but one potential explanation is the increased activity of NAD⁺-dependent enzymes such as sirtuins which can influence the processes involved in aging biology, such as chromatin modelling, DNA methylation and mitochondrial function. Taken together, these findings highlight the potential of epigenetic clocks as biomarkers possibly capturing subtle cellular adaptations not reflected in standard clinical readouts, and also raise the possibility that NR may promote muscle tissue health and potentially affect aging-related pathways. Further studies are warranted to determine whether these epigenetic shifts translate into functional or clinical benefits in humans over time, and to establish optimal NR supplementation conditions to beneficially influence metabolic outcomes and biological aging.

Epigenetic age is affected by an individual’s genome as well as environmental and lifestyle factors (Reynolds et al. 2020), such as smoking (Ramirez et al. 2025; Klopack et al. 2022). The inclusion of monozygotic twins, who share their genomic sequence, in the NR trial, allowed us to explore the relative contributions of individual-level effects versus shared genetic and environmental influences on the variability of the EAA. The ICC analyses, which describe the similarity in a parameter within-pair versus between-pair, demonstrated that co-twins were highly similar in most of their EAA measures at baseline, but differed more in how their EAA changed during NR supplementation. This suggests that the response to NR is primarily driven by individual-specific factors, such as lifestyle effects or random variation, rather than shared genetic or environmental influences. Interestingly, for clocks such as MEAT and PC-Horvath, which incorporate muscle-derived methylation data in their development, we observed relatively high ICC for the change in skeletal muscle EAA after NR intervention. This similarity between the co-twins (even slightly higher than at baseline) indicates that, for these clocks, the NR-induced change in EAA is partially accounted for by genetic and shared environmental factors. In line, we have previously also shown that monozygotic co-twins did not differ from each other in their response to NR in other epigenetic measures, global DNA methylation and mitochondrial-related CpG methylation in muscle (Lapatto et al. 2023). A previous study has suggested that NR has more effect on EAA in those individuals with higher levels of inflammation (Norheim et al. 2024). Therefore, our findings and prior research indicate complex interplay between genetic and disease- or lifestyle-related factors in NR supplementation responses. Defining these factors and quantifying their relative contributions is essential, yet challenging.

Some studies have reported that NR supplementation can increase mitochondrial content in mouse muscle (Zhang et al. 2016; Frederick et al. 2016) and cultured human hepatocytes (Lee & Yang 2019). However, when we mimicked the chonic effect of NR in human myotubes, 3-day NR treatment significantly increased intracellular NAD⁺ levels, but this elevation did not translate into changes in mtDNAq or EAA measures, possibly due to the insufficient duration of the NR addition. Alternatively, it may be that direct addition of NR to myotubes have different effects than systemic administration, where interactions with other cell types are present and may be required for full mitochondrial responses. These findings suggest that the effects of NR on epigenetic aging may depend on systemic or tissue-level interactions that are absent in isolated cell cultures and that short treatment durations *in vitro* may be insufficient to elicit measurable changes. Furthermore, the epigenetic clocks used in this study were developed using bulk tissue samples composed of multiple cell types, which may limit their applicability to homogeneous cell populations like cultured myotubes, particularly if the observed effects are primarily driven by other cell types. Other biological age estimators such as muscle-specific protein clocks (Goeminne et al. 2025) or other aging biomarker panels (Hartmann et al. 2023) may provide alternative insights into the effects of NR both *in vivo* and *in vitro*.

Exercise, including HIIT, is well known to confer numerous health benefits (Cartee et al. 2016; Mølmen et al. 2025; Gleeson et al. 2011; Robinson et al. 2017), and influence skeletal muscle DNA methylation patterns (Voisin et al. 2024) and epigentic aging markers (Ostaíza et al. 2025; Jones III et al. 2023) to represent a more youthful methylome. In contrast, by using epigenetic clocks, we observed that HIIT was mainly associated with increased EAA, in both 4-week Gene SMART and 6-week EpiH studies when measured with DunedinPACE. This discrepancy with previous finding could reflect transient molecular stress responses rather than long-term adaptations, as tissue sampling 48-72 hours after the last intervention may potentially capture the acute remodeling, inflammation, increased mitochondrial reactive oxygen species production or shifts in cellular turnover (Peake et al. 2016) that temporaily accelarate aging signals. Epigenetic clocks are sensitive to dynamic changes and may not always align with long-term health outcomes. It is also possible that tissue-level modifications induced by exercise, such as shifts in skeletal muscle cell type composition (Lovrić et al. 2022), contribute to the observed changes in epigenetic ages. For instance, the EpiH DNA methylation was measured from extracted muscle fibers as opposed to bulk muscle tissue in Gene SMART which may partially explain some of the different results between the two studies. Furthermore, the effects of HIIT were generally opposite to those observed after NR supplementation. Clocks that showed decreased EAA after NR supplementation—such as DunedinPACE—tended to show increased EAA following HIIT, while others like PCGrimAge showed the reverse pattern. This divergence suggests that NR and HIIT may influence distinct epigenetic pathways or reflect differences in the duration and nature of the interventions. However, unlike in the NR trial where blood NAD^⁺^ levels increased post-supplementation (Lapatto et al. 2023), we did not assess blood or muscle NAD^⁺^ post-exercise. Thus, whether EAA changes with NR are driven by increased NAD^⁺^ availability, or occurs similarly in the HIIT studies, remains unclear. Further research is warranted to determine whether these early changes in EAA represent adaptive remodeling or potential stress-induced aging acceleration, and how they relate to long-term health and longevity.

Increased mitochondrial content and activity been generally associated with lower EAA measures in blood (Kabacik et al. 2022), as evidenced by clocks like DunedinPACE (Wang et al. 2025). In contrast, we observed a positive correlation between changes in skeletal muscle mtDNAq and MEAT EAA, indicating that individuals with greater increases in mtDNAq after NR tended to exhibit smaller reductions in EAA. Similarly, following HIIT, MEAT-derived EAA was positively correlated with changes in CS activity, another marker of mitochondrial content. Consistent patterns across both interventions suggests that the MEAT clock may reflect molecular alterations in muscle. One potential explanation is that increases in mtDNA and CS activity may indicate higher cellular stress (Lee et al. 2000) which could be linked to elevated EAA. Alternatively, these relationships may be influenced by other factors such as inflammation, which can affect both EAA and mitochondrial content (Franceschi et al. 2018). Overall, these findings suggest that epigenetic aging in muscle is linked to cellular stress and metabolic changes, but cannot be explained by mitochondrial changes alone, highlighting the complex biology behind epigenetic aging.

Epigenetic clocks developed for blood may not fully capture aging-related changes in other tissues (Apsley et al. 2025). While we previously showed that blood-based clocks are weaker in detecting relevant changes in muscle aging and show weak associations with physical function (Sillanpää et al. 2021), we aimed to examine how these clocks respond to different interventions within the same individual and evaluate their applicability. Blood-developed clocks often showed high deviations from chronological age when calculated from muscle but were moderately to highly correlated with age, suggesting that although some CpGs may not reflect muscle-specific aging, these clocks still capture relevant age-related methylation signatures shared across tissues. In contrast, the MEAT clock—designed for muscle—demonstrated superior accuracy and responsiveness, reinforcing the importance of tissue-targeted clocks in aging research. Furthermore, we observed no cross-tissue correlations between clocks within individuals after NR, even for those showing significant decreases in both tissues such as DunedinPACE, which further unferscores the tissue- and clock-specific nature of epigenetic aging. Future work should examine the epigenetic signatures driving these clock-specific changes after NR and HIIT and their links to underlying biological processes.

This study has several limitations that should be considered when interpreting the findings. First, modest sample sizes may limit statistical power and generalizability, particularly given random biological and technical variation in EAA estimates. Second, the NR and HIIT interventions lacked placebo controls, making it difficult to rule out confounding effects or natural fluctuations in EAA over time. Third, heterogeneity existed across studies in intervention lengths, participant characteristics, and mitochondrial measurement methodologies, likely contributing to discrepancies. Differences in methylation array versions (EPIC v1 vs. v2) may also have influenced epigenetic ages. Lastly, although we aimed to mimic chronic NR treatment in human myotubes, experimental conditions differed substantially from *in vivo* situation. Despite these challenges, data from controlled human interventions with access to the primary target tissue—rather than blood-based surrogates only—offers unique and robust perspective into tissue-specific epigenetic aging in response to NR supplementation and HIIT.

Our findings provide novel insights by demonstrating that both NR supplementation and exercise can modulate epigenetic aging in muscle, and that these effects appear to be both clock- and intervention-specific. These results lay the groundwork for future research aimed at disentangling the molecular mechanisms underlying fluctuations in human biological aging in response to mitochondria targeting interventions, ideally through studies incorporating placebo controls and longitudinal designs. Furthermore, integrating mechanistic analyses will be essential to determine if and how changes in epigenetic age relate to mitochondrial function, NAD⁺ metabolism, and ultimately, age-related functional decline and disease risk.

## Funding

The study was supported by the Research Council of Finland (335445, 314455 for EP, and 328685, 307339, 297908, 251316 for MO, 272376, 266286, 314383, 335443, 369181 for KHP, 361956, 338417 for SH, 336823, 352792 for JK), Research Council of Finland Profi6 funding (336449) awarded to the University of Oulu (EP), Finnish Diabetes Research Foundation (EP, SH, KHP), the Minerva Foundation, the Liv och Hälsa sr., the Sigrid Juselius Foundation (MO, KHP), The Finnish Cultural Foundation Kymenlaakso Fund (MO and AH), Orion Foundation (SH, BWK), Finnish Medical Foundation (KHP, SH), Finnish Foundation for Cardiovascular Research (KHP), Novo Nordisk Foundation (NNF10OC1013354, NNF17OC0027232, NNF20OC0060547, NNF24OC0091683, NNF25SA0103783 for KHP; NNF23SA0083953 and NNF25OC0100827 for SH; NNF24SA0090438 for BWK), Paulo Foundation (SH), Gyllenberg Foundation (KHP), Paavo Nurmi Foundation (SH), Helsinki University Hospital Research Funds (SH, KHP), Government Research Funds (KHP, SH), University of Helsinki (KHP), Juho Vainio Foundation (SA) and Yrjö Jahnsson Foundation (BK).

## Ethics statement

The NR trial study protocol was approved by the Ethics Committee of the Helsinki University Central Hospital (protocol number 270/13/01/2008). The EpiH study was approved by the Ethical Committee of Copenhagen (H-3-2012-024). The Gene SMART study was approved by the Victoria University Human Ethics Committee (HRE13-223). Written informed consent was obtained from all participants of each study.

## Data availability

The FTC DNA methylation is part of the ‘Twin Study’ and deposited with the Biobank of the Finnish Institute for Health and Welfare [https://thl.fi/en/research-and-development/thl-biobank/for-researchers/sample-collections/twin-study]. For details on accessing the data, see: https://thl.fi/en/research-and-development/thl-biobank/for-researchers/application-process. The EpiH DNA methylation data will be deposited into a public repository. In the meantime, the data can be requested from the corresponding author. The Gene SMART DNA methylation data is deposited to Gene Expression Omnibus with accession code **<u>GSE171140</u>**.

## Author contributions

A.H. and M.O. conceptualized and designed the study. A.H. preprocessed the data, performed statistical analyses, visualized the results, and wrote the first draft of the manuscript. R.K. and E.P. contributed to the conceptualization of the myotube cell culture experiment. L.U.K. conducted the myotube cell culture experiment. S.A. contributed to the interpretation of findings and offered theoretical guidance on exercise research. J.K., B.W.K., S.H., K.H.P., and E.P. conceived, collected, and generated data for the NR trial study. I.B., J.W.H., L.G., R.S., and S.L. conceived, collected, and generated data for the EpiH study. M.J., R.G., and N.E. conceived, collected, and generated data for the Gene SMART study. All authors provided critical feedback on the manuscript and approved the final version for submission.

## Conflict of interest

The authors have no conflicts of interest to declare.

## Notes

### Competing Interest Statement

The authors have declared no competing interest.

### Clinical Trial

NCT03951285

### Author Declarations

The Ethics Committee of the Helsinki University Central Hospital (protocol number 270/13/01/2008) gave ethical approval for the NR trial. The Ethical Committee of Copenhagen (H-3-2012-024) gave ethical approval for the EpiH study. The Victoria University Human Ethics Committee (HRE13-223) gave ethical approval for the Gene SMART study. Written informed consent was obtained from all participants of each study.

