## Supplementary Materials for "The divergent effects of nicotinamide riboside and high-intensity exercise training on skeletal muscle epigenetic aging"

### Supplementary Tables

**Supplementary Table 1.** Person correlation coefficients (R2) and mean absolute error (MAE) between epigenetic ages and chronological ages across all human cohorts in the study.

| | NR trial | | | | EpiH | | Gene SMART | |
|---|---|---|---|---|---|---|---|---|
| | Blood | | Muscle | | Muscle | | Muscle | |
| | R2 | MAE | R2 | MAE | R2 | MAE | R2 | MAE |
| PCHorvath | 0.902 | 7.201 | 0.719 | 9.894 | 0.709 | 13.006 | 0.540 | 15.189 |
| PCHannum | 0.929 | 10.690 | 0.717 | 14.577 | 0.721 | 13.570 | 0.562 | 20.925 |
| PCPhenoAge | 0.801 | 4.599 | 0.690 | 30.356 | 0.784 | 20.506 | 0.530 | 35.232 |
| PCGrimAge | 0.920 | 13.636 | 0.976 | 27.474 | 0.993 | 24.847 | 0.966 | 30.557 |
| GrimAgev2 | 0.813 | 8.455 | 0.956 | 6.545 | 0.980 | 5.894 | 0.903 | 10.492 |
| MEAT* | - | - | 0.937 | 2.618 | 0.969 | 3.255 | 0.880 | 3.582 |

* MEATv2 was used in all other cohorts except for Gene SMART

**Supplementary Table 2. Epigenetic age acceleration measures at baseline and after 5-month NR supplementation (n=36 individuals), along with intra-class correlation coefficients (ICCs) (n=15 complete twin pairs) in blood.** P-values are derived from linear mixed-effects models adjusted for chronological age, sex, smoking, BMI, with personID nested within familyID as a random effect. P-value<0.05 are bolded.

| **Clock** | **Mean (SD)* at baseline** | **Mean (SD)* at 5-month** | **Change (SD)*** | **P-value** | **ICC (95% CI) baseline** | **ICC (95% CI) change** |
|---|---|---|---|---|---|---|
| DunedinPACE | 0.96 (0.10) | 0.94 (0.09) | -0.02 (0.05) | **0.050** | 0.55 (0.11-0.83) | - |
| PCHorvath | 0.51 (2.86) | -0.58 (2.92) | -1.09 (1.67) | **3.0E-04** | 0.86 (0.63-0.95) | 0.26 (0-0.66) |
| PCHannum | 0.31 (2.46) | -0.37 (2.76) | -0.68 (1.38) | **0.011** | 0.64 (0.21-0.86) | 0.42 (0-0.76) |
| PCPhenoAge | -0.15 (5.04) | -0.20 (5.20) | -0.05 (2.79) | 0.865 | 0.60 (0.14-0.85) | 0.69 (0.27-0.89) |
| PCGrimAge | -0.24 (2.59) | 0.00 (2.86) | 0.24 (1.24) | 0.291 | 0.33 (0-0.68) | 0.28 (0-0.68) |
| GrimAge2 | -0.27 (4.10) | 0.02 (4.31) | 0.29 (1.92) | 0.401 | 0.48 (0-0.78) | 0.13 (0-0.6) |

* Units are in years, except for DunedinPACE, which is expressed in years/year.

**Supplementary Table 3. Differences in NR-induced changes in epigenetic age acceleration (EAA) in BMI-discordant monozygotic twin pairs, comparing leaner co-**

**twins to their heavier counterparts (reference group) in muscle (n = 14 pairs) and blood (n= 15 pairs).** Only clocks that showed a significant main effect of NR are shown.

| Tissue | Clock | Coefficient | P-value |
|---|---|---|---|
| Muscle | DunedinPACE | 0.012 | 0.664 |
| | PCHannum | 0.610 | 0.167 |
| | PCGrimAge | 0.176 | 0.675 |
| | MEAT | -0.060 | 0.936 |
| WBC | DunedinPACE | -0.014 | 0.413 |
| | PCHannum | -0.361 | 0.381 |
| | PCHorvath | -0.416 | 0.379 |

**Supplementary Table 4. Correlation between changes in epigenetic age acceleration (EAA), and citrate synthase (CS) activity and $VO_{2max}$ after a 6-week high-intensity interval training (HIIT) intervention, stratified by younger (n = 19; age 21–42 years) and older (n = 20; age 55–74 years) participants in the EpiH cohort.**

| | EAA | Cor (old) | Cor (young) | P.value (old) | P.value (old) |
|---|---|---|---|---|---|
| CS activity | DunedinPACE | 0.087 | -0.157 | 0.750 | 0.575 |
| | GrimAge2 | 0.033 | -0.110 | 0.902 | 0.697 |
| | MEAT | 0.223 | 0.294 | 0.407 | 0.287 |
| | PCGrimAge | 0.128 | -0.579 | 0.636 | **0.024** |
| | PCHannum | -0.179 | -0.454 | 0.507 | 0.089 |
| | PCHorvath | -0.252 | -0.466 | 0.347 | 0.080 |
| | PCPhenoAge | -0.275 | -0.347 | 0.302 | 0.205 |
| $VO_{2max}$ | DunedinPACE | 0.194 | 0.246 | 0.426 | 0.296 |
| | GrimAge2 | -0.457 | -0.034 | 0.049 | 0.885 |
| | MEAT | -0.413 | -0.414 | 0.078 | 0.070 |
| | PCGrimAge | 0.155 | -0.047 | 0.526 | 0.844 |
| | PCHannum | 0.268 | 0.017 | 0.267 | 0.942 |
| | PCHorvath | 0.112 | 0.338 | 0.647 | 0.145 |
| | PCPhenoAge | 0.059 | 0.129 | 0.811 | 0.587 |

### Supplementary Figures

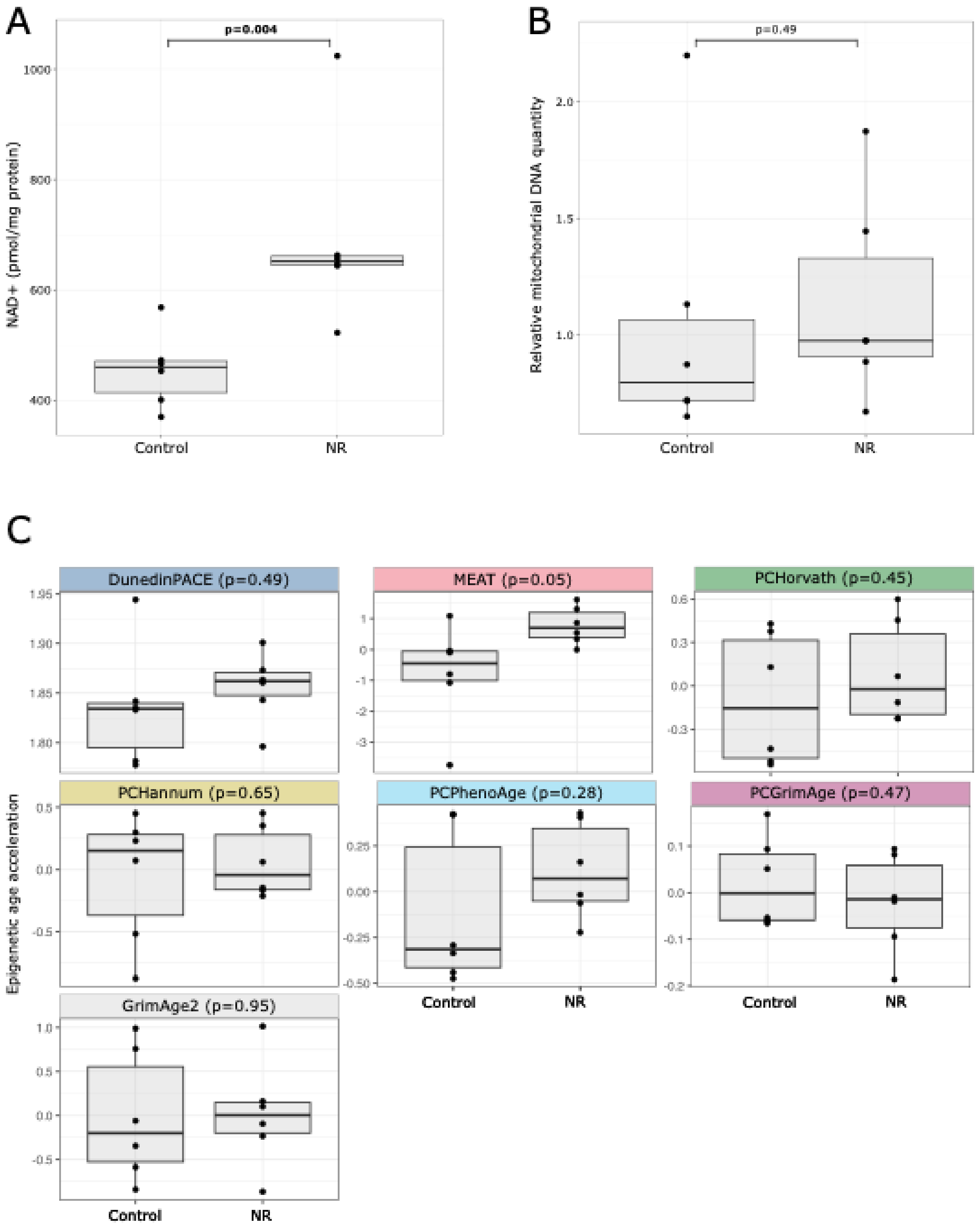


**Supplementary Figure 1.** A) Levels of NAD+, B) mitochondrial DNA quantity (mtDNAq) and C) epigenetic age acceleration measures in control and NR-treated cells. P-values are derived from Wilcoxon signed-rank tests (A,B) or from linear models (C).

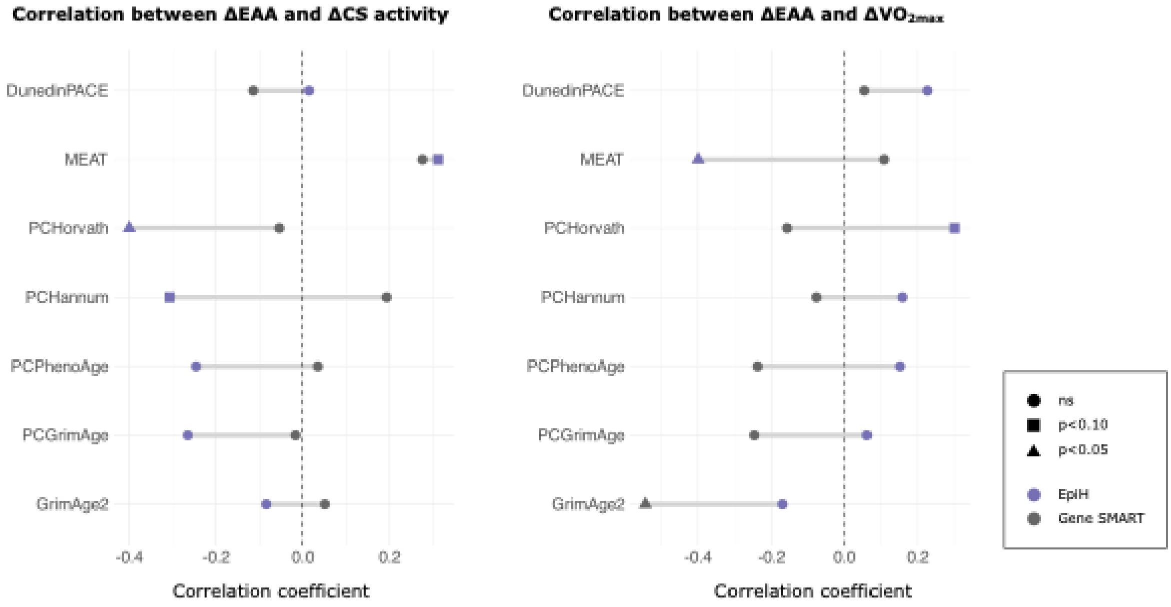


**Supplementary Figure 2.** Correlations between changes in EAA and changes in citrate synthase (CS) activity and cardiorespiratory fitness ($VO_{2max}$) following HIIT in the EpiH and Gene SMART cohorts. Each point represents a correlation coefficient for one cohort, with grey lines connecting paired estimates to illustrate differences between cohorts. Point shapes indicate significance levels based on p-values.
Δ = post value vs. pre value, ns = not significant

### Supplementary Methods

<u>Myotube cell culture</u>

Myoblasts were cultured in high-glucose DMEM (Gibco, 10313-021) supplemented with 20% FBS (Gibco, A5256701), 2 mM L-glutamine (Gibco, 51500-056), 100 U/mL penicillin, and 100 µg/mL streptomycin (Gibco, 15140-122) and differentiated to myotubes by changing the media to high-glucose DMEM supplemented with 2% horse serum (Gibco, 26050-088), 1% Insulin-Transferrin-Selenium-Ethanolamine (ITS-X) supplement (Gibco, 51500-056), 2 mM L-glutamine, 100 U/mL penicillin and 100 ug/mL streptomycin.
